# Environment influences SARS-CoV-2 transmission in the absence of non-pharmaceutical interventions

**DOI:** 10.1101/2020.09.12.20193250

**Authors:** Thomas P. Smith, Seth Flaxman, Amanda S. Gallinat, Sylvia P. Kinosian, Michael Stemkovski, H. Juliette T. Unwin, Oliver J. Watson, Charles Whittaker, Lorenzo Cattarino, Ilaria Dorigatti, Michael Tristem, William D. Pearse

## Abstract

As COVID-19 continues to spread across the world, it is increasingly important to understand the factors that influence its transmission. Seasonal variation driven by responses to changing environment has been shown to affect the transmission intensity of several coronaviruses. However, the impact of the environment on SARS-CoV-2 remains largely unknown, and thus seasonal variation remains a source of uncertainty in forecasts of SARS-CoV-2 transmission. Here we address this issue by assessing the association of temperature, humidity, UV radiation, and population density with estimates of transmission rate (*R*). Using data from the United States of America, we explore correlates of transmission across USA states using comparative regression and integrative epidemiological modelling. We find that policy intervention (‘lockdown’) and reductions in individuals’ mobility are the major predictors of SARS-CoV-2 transmission rates, but in their absence lower temperatures and higher population densities are correlated with increased SARS-CoV-2 transmission. Our results show that summer weather cannot be considered a substitute for mitigation policies, but that lower autumn and winter temperatures may lead to an increase in transmission intensity in the absence of policy interventions or behavioural changes. We outline how this information may improve the forecasting of COVID-19, its future seasonal dynamics, and inform intervention policies.

## Introduction

In late 2019 a novel coronavirus originating in Wuhan City (Hubei, China)^1^ began to rapidly spread through the human population. Since March 2020 this disease, COVID-19, has been recognised as a global pandemic by the World Health Organisation^2^. The causative agent, SARS-CoV-2, is a close relative of the 2003 SARS coronavirus^1^, although it appears to have several differences including a higher basic reproduction number^3^ (*R*_0_; the average number of people infected by a carrier at the onset of an epidemic). Understanding the factors influencing SARS-CoV-2 transmission is key for understanding the current patterns of transmission and for refining predictions of the future spread of SARS-CoV-2. Other coronaviruses display seasonal cycles of transmission and up to 30% of seasonal ‘colds’ are caused by coronaviruses^4^. Thus, as many Northern-hemisphere countries relax the non-pharmaceutical interventions initially imposed to control COVID-19, there is a pressing need to understand whether seasonality will enhance or drive a ‘second wave’ of COVID-19 outbreaks as they move into autumn or winter^5^.

SARS-CoV-2 is an enveloped RNA virus which is structurally (if not phylogenetically) similar to other RNA viruses such as influenza, MERS, and HcoV-NL63^6^, that are known to display seasonal dynamics due to their physical properties. For example, high temperatures and low humidity can have a negative effect on influenza transmission by reducing the efficiency of respiratory droplet transmission^7,8^. Similar effects are seen in transmission of coronaviruses^9-11^, where high environmental temperatures break down viral lipid layers to inactivate virus particles that are in the air or deposited on surfaces^10,12^. However, assessing the role of environment during a disease outbreak is challenging^13^ because human factors such as population density, herd immunity, and behaviour are likely the main drivers of transmission^14-16^. Moreover, the non-pharmaceutical control measures and behavioural changes in response to COVID-19 have been unprecedented in the modern era. These difficulties have hindered the quantification of the impact of environment on SARS-CoV-2 transmission, making it harder to generalise and synthesise observations across regions with their differing climates. Despite these caveats, various early studies have already reported effects of environmental variables such as temperature, humidity, UV levels, and wind-speed on the transmission of SARS-CoV-2^17-24^. While, in general, most studies appear to support increased transmission rates under cool, dry conditions^18^, conflicting results have been observed^21,25^ and collectively the environmental signal appears to be weak^5^. Much of the variability in these early results is likely due to the use of inappropriate response variables (such as cases or fatalities) which fail to capture the intrinsic variations in transmission intensity driven by the effects of non-pharmaceutical intervention measures^5^. Furthermore, COVID-19 has taken hold in many places with diverse climates and there are obvious examples of high transmission rates under hot, humid conditions, *e.g*. in Brazil^26^.

Accurate assessment of the role environmental factors have played so far in the spread of SARS-CoV-2 may provide insight into the future seasonality of the disease. This is because seasonal outbreaks of viruses are often driven by their responses to favourable (seasonal) changes in weather^27^. Most epidemiological forecasts make use of some variant of the Susceptible-Infected-Recovered (SIR) framework and/or focus on the impacts of government-level mitigation (*e.g*. Kissler et al.^28^, Walker et al.^29^). Few epidemiological models incorporate environmental impacts and, when they do, they assume COVID-19 responds in a manner identical to related coronaviruses because we lack data on SARS-CoV-2’s environmental (and thus seasonal) responses (*e.g*. Baker et al.^18^). This is despite theoretical demonstrations of the potential role of environment in driving future seasonality of SARS-CoV-2^22,30^ and the empirical evidence in structurally-similar viruses outlined above. Efforts to incorporate climate into COVID-19 forecasting have focused on regression-type models of cases and fatalities (*e.g*. Araujo and Naimi^17^), which are unreliable when diseases are in the growth/expansion phase^31^. Furthermore, such models conflate environmental controls on occurrence with other drivers such as public health interventions (*e.g*., the effects of lockdown measures to contain the pandemic)^31^ as both are changing similarly through time. Such models are unlikely to yield useful insights and may be misleading to policymakers^13^. To address this knowledge gap, there is a need for a true synthesis of environmental modelling with well-established epidemiological approaches.

Here we investigate the role of environment in the transmission of SARS-CoV-2 by incorporating environmental factors into an existing epidemiological framework that has been applied globally^32-34^, and to the USA in particular^35^. The USA is a large country with great variation in climate across which case and policy intervention data are comparable, permitting us to disentangle the role of environmental drivers in SARS-CoV-2 transmission. We begin by exploring associations between the environment (temperature, humidity, UV radiation, and population density) and transmission intensity independently estimated before and during stay-at-home orders (henceforth termed “lockdown”). We used the basic reproduction number (*R*_0_) for our pre-lockdown transmission intensity estimates, and the time-varying reproduction number (*R_t_*, the reproduction number, *R*, at a given time, *t*) averaged across an appropriate time-window for our during-lockdown estimates. After confirming a potential role for the environment, we verify and more accurately quantify the relative roles of temperature and population density by integrating them into an existing semi-mechanistic epidemiological framework^35^. While we find strong evidence that temperature and population density are associated with SARS-CoV-2 transmission, we emphasise that our findings also re-confirm that the major drivers of transmission rates are public policy and individual behaviour. Through our use of existing, robust sources of forecasts and models, our findings can be easily incorporated into workflows already used by policymakers, as we detail here.

## Results

When analysed jointly, the *R*_0_ of all USA states are fairly well predicted by all explanatory variables included in the regression model *(i.e*. population density, temperature, absolute humidity and UV radiation), with an overall model *adjusted r*^2^ of 58% (supplementary table S1). However, UV radiation is a very weak predictor of *R*_0_, while temperature and absolute humidity show sufficiently strong correlations with each other (*r* = 0.85) that we cannot disentangle their contributions to *R*_0_ due to high inflation of variances (supplementary table S1). This is further demonstrated through principal components analysis, where temperature and absolute humidity fall along the same principal component axis (supplementary figure S1). We therefore focused on temperature as the best fitting climate variable (assessed by Pearson’s *r*, supplementary table S2).

When regressed against temperature and log_10_-transformed population density only, we find that *R*_0_ significantly increases with population density and decreases with temperature (fig. 1; both *p* < 0.001, table 1). We see a stark difference, however, when analysing *R_t_* during lockdown (defined as the mean *R_t_* recorded over the 14 day period following a stay at home order): much less of the variation in *R_t_* is explained by the regression model (*adjusted r*^2^ = 18%), vastly lower model coefficients for explanatory variables (*i.e*., much lesser correlations; supplementary table S3, but note that population density is still a significant predictor), and much lower *R_t_* estimates overall (paired *t*_39_ = 21.1; *p* < 0.001; figure 1b). Additionally, if we regress the combined *R*_0_ and *R_t_* estimates against temperature and population density, using lockdown as a binary interaction term, we find a significant interaction between lockdown and temperature (*p* < 0.001, supplementary table S5), *i.e*. lockdown mediates the effects of climate on transmission.

**Figure 1:**
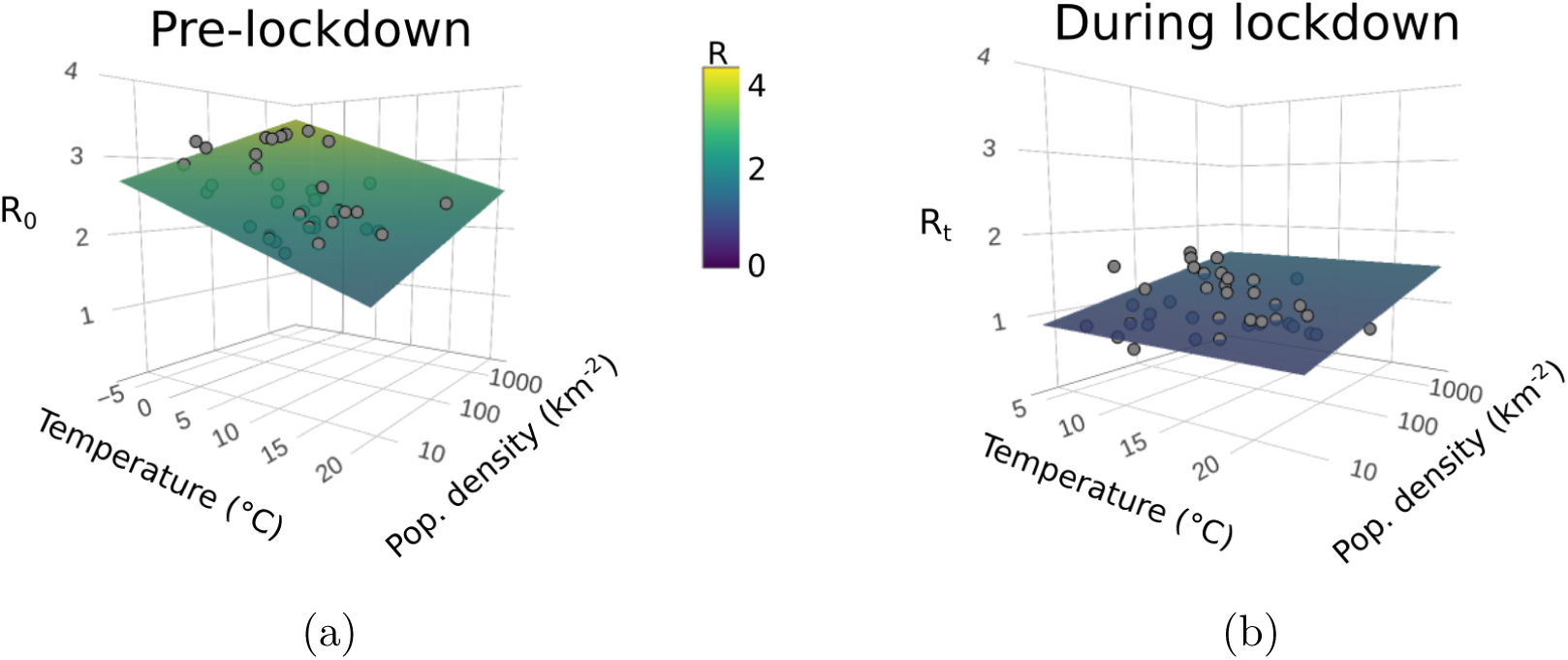
*R*_0_ is affected by the environment, but the impact of lockdown is greater. **(a.)** *R*_0_ plotted against temperature (averaged across the two weeks prior to the *R*_0_ estimate) and log_10_-transformed population density (people per km^2^) for each USA state (grey points). Surface shows the predicted *R*_0_ from the regression model (table 1). Temperature has a negative effect on *R*_0_ at state-level in the USA, whilst population density has a positive effect (table 1). **(b.)** The mean *R_t_* for the two weeks following a state-wide stay-at-home mandate (*i.e*., during lockdown) plotted against average daily temperature for the same period and log_10_-transformed population density. The effects of temperature and population density are much weaker in the mobility-restricted data and R is reduced overall. The same colour scale, given in the centre of the figure, is used across both sub-plots.

**Table 1:** Population density and temperature are drivers of *R*_0_ at state-level in the USA. Multiple *r*^2^ = 0.6037, *adjusted r*^2^ = 0.5822, F_2,37_ = 28.18, *p* < 0.001. Scaled estimates are coefficients when predictors are scaled to have mean = 0 and SD = 1. Scaling our explanatory variables means our coefficients are measures of the relative importance of each variable. In contrast to our epidemiological modelling, temperature is a greater driver of pre-lockdown *R*_0_ than population density (log_10_-transformed). * = *p* < 0.05

| | Scaled Estimate | Std. Error | $t$ value | $p$-value |
| --- | --- | --- | --- | --- |
| (Intercept) | 2.5600 | 0.0494 | 51.80 | < 0.001* |
| Temperature | -0.2801 | 0.0516 | -5.43 | < 0.001* |
| $\log_{10}$(Pop density) | 0.1921 | 0.0516 | 3.72 | < 0.001* |

The strong correlates of population density and temperature on *R*_0_ across the United States were echoed in our climate-driven Bayesian modelling of daily variation in R_t_. Posterior medians of the scaled coefficients of (log_10_-transformed) population density and daily temperature were 0.68 and -0.48, respectively. These coefficients were strongly supported (both Bayesian probabilities > 99.9%), and suggest that greater population density is approximately 1.4 times a greater driver of higher transmission than colder temperature (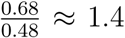). Changes in mobility (such as those induced by stay-at-home measures) have the potential to mitigate these impacts of both population density and temperature (figure 3). Our model suggests that even quite large variation in underlying transmission driven by either variation in temperature through time, or in population density across space, can be mitigated by reductions in mobility (see also supplementary figure S2). Critically, however, the posterior distributions are skewed, particularly for population density: high population density may be difficult to mitigate except through large mobility reductions (as shown by the credibility intervals in figure 3). We emphasise that other transmission mitigation decisions, such as hand-washing, mask-wearing, and physical distancing, were not assessed in our model. We highlight that the posterior estimates of environment and average mobility were correlated (Pearson’s *r* = 0.30 for temperature and *r* = -0.32 for population density). This likely results from correlated changes in mobility and temperature through time, and makes the estimated mobility reductions in figure 3 conservative (*i.e*., we potentially report larger mobility reductions than would be necessary to mitigate environmental effects).

## Discussion

Here, by combining epidemiological models and outputs with spatial climate data, we show that environment (specifically cold, but also the correlated low-humidity conditions) can enhance SARS-CoV-2 transmission across the USA. Critically, however, these environmental impacts are weaker than that of population density which is, itself, a weaker driver than policy intervention (*i.e*., lockdown). Below, we suggest that the accuracy of forecasts of SARS-CoV-2 transmission, in particular across seasons, could be improved by incorporating temperature, as well as population density, in a robust, reproducible manner as we have done here.

### The role that environment plays in transmission

Across these state-level USA data, we found a significant negative effect of temperature on SARS-CoV-2’s *R*_0_ and a significant positive effect of population density. An important caveat to this, however, is the collinearity between temperature, absolute humidity, and to a lesser degree, UV levels. The strong correlations between these environmental drivers mean that we are unable to discern the effects of each in a single model and therefore we focus on temperature as the most reliable environmental predictor. After accounting for the effect of population density on transmission (table 1), temperature’s effect is striking (figure 2). We also tested the effects of our predictor variables on *R_t_* for times where strict lockdown measures were in place. When these mobility restrictions are in place, we observe no significant effects of temperature on *R_t_*, *i.e*. the effects of lockdown dampen any environmental effects so as to make them inconsequential (figure 1b; supplementary table S3). Furthermore, under lockdown conditions the overall transmission rates are vastly reduced. Through our epidemiological modelling approach we are able to account for these effects (as mobility changes are explicitly incorporated), and find that higher population densities and lower temperatures drive increased R_t_. Moreover, the formulation of our epidemiological model ensures that under high mobility reductions, changes in environment have little effect on *R_t_*, mirroring our regression findings (see Methods and supplementary figure S2).

**Figure 2:**
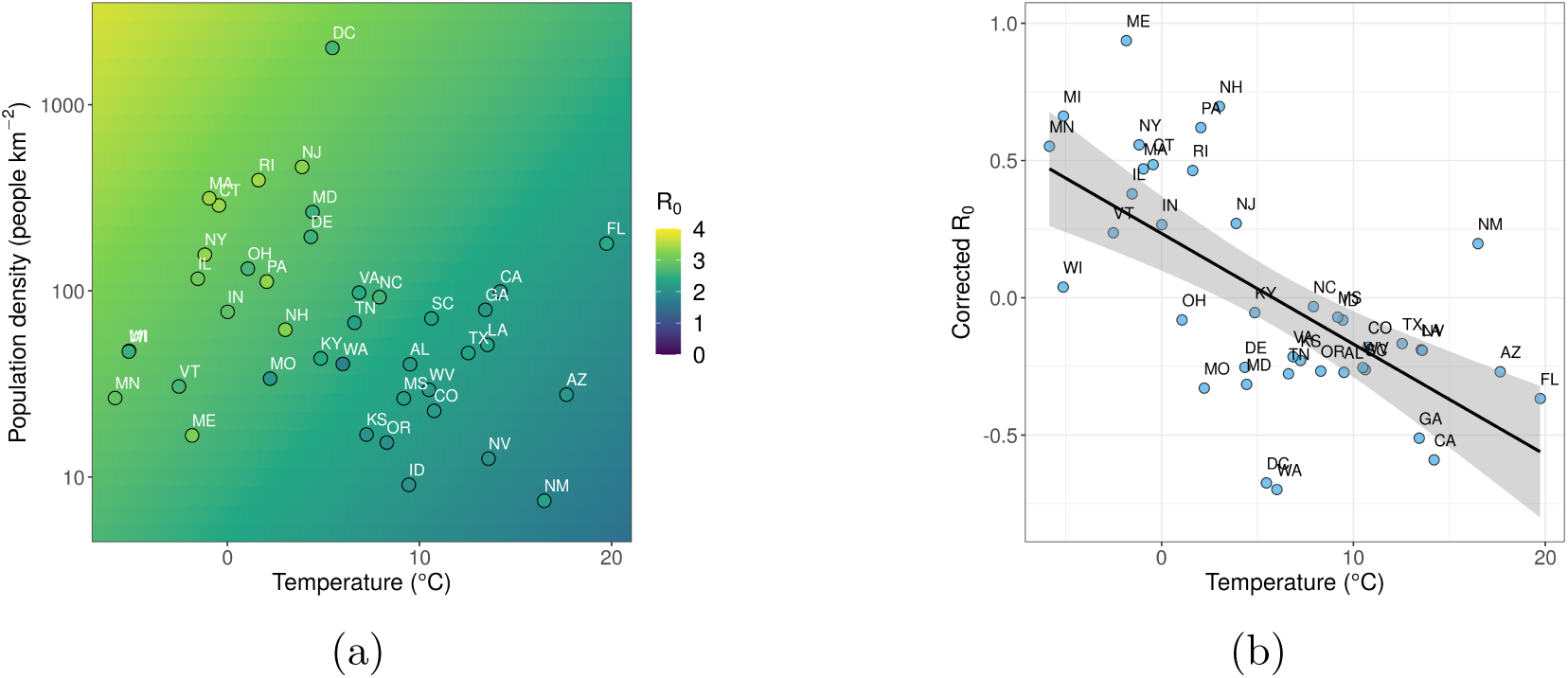
The relative importance of temperature and population density as drivers of pre-lockdown *R*_0_. **(a.)** Heatmap of the regression model *R*_0_ predictions, with USA state-level *R*_0_ point estimates overlaid. High population densities and low temperatures drive increases in SARS-CoV-2 *R*_0_. This is a 2D representation of the regression plane in fig. 1a, using the same colour-scale. **(b.)** Residuals from a linear regression of *R*_0_ against log_10_-transformed population density (“Corrected *R*_0_”), plotted against temperature. This illustrates that, when considering population density alone, Ro is overestimated in cold states and underestimated in warm states. After accounting for population density, there is a significant effect of temperature upon *R*_0_ (see table 1). In both figures, points are highlighted with standard two-letter state codes; MN and FL refer to Minnesota and Florida, respectively, and are referred to in the discussion.

The precise physiological mechanisms for temperature-dependant inactivation in SARS-CoV-2 are still not known, but animal models for influenza have shown that increased viral transmission at lower temperatures can be due to effects on the host^7,8^. In animal models, this is proposed to be due to the combined effects of higher titres of viral particle shedding and greater viral stability in nasal passages of those housed in cooler conditions^8^. In addition to host effects, the persistence time of the virus outside of the body is expected to be negatively affected by higher environmental temperatures, which cause viral inactivation via breakdown of their lipid layers^10,12^. We emphasise that both the direct host effects, and the potential effects of environment on viral stability, are likely moderated (if not mitigated) by indoor heating^37^, although the same may not always be true of humidity. Contact rate is related to population density^16^, and so it is unsurprising that population density was a significant factor in our analysis (figure 1a). We stress that temperature was not a driver of transmission under lockdown and the effects of population density were lessened (figure 1b): outdoor conditions and population density matter little for indoor transmission.

There are important methodological caveats to our findings. Dynamics and reporting between USA states are known to be variable^38^, introducing a level of uncertainty to our findings. Furthermore, lockdown measures were (and continue to be) quite heterogeneous across the USA, with different states displaying different levels of response to COVID-19^39^. Through our epidemiological modelling approach we are able to account for these different state-level responses using google mobility data. We can also observe other potential confounding factors in these analyses. Across the USA, the north-eastern states in the vicinity of the major transport hub of New York City (*e.g*., NY, NJ, ME, PA, RI, and CT) tend to have generally higher *R*_0_ than predicted, whilst west-coast states (*e.g*., WA, CA, and OR) have lower *R*_0_ than predicted (fig 2a). While this type of effect could be due to preemptive protective measures taken by states prior to COVID-19 outbreaks, we likely mitigated this by removing states that initiated non-pharmaceutical interventions before our first time-step (see methods). A further confounding factor may be seen if temperature affects human behaviour, thus making it difficult to disentangle the effects of climate from changes to mobility. We do find a link between the average mobility and temperature coefficients in our Bayesian modelling, suggesting a degree of collinearity, however (perhaps surprisingly), we see no direct correlations between daily temperature and recreational mobility trends for parks (see supplementary information). Again, this highlights the importance of human behaviour as a confounding factor in analyses of environmental drivers on SARS-CoV-2 transmission. Future work should consider finer-grained population density, as well as the presence of major transport hubs in a given region.

### Policy relevance of our findings

Our results comparing SARS-CoV-2 transmission rate before and during lockdown support the idea that the major driver of transmission is public health policy ^32,40,41^ (see figure 1). Once stay-at-home measures were implemented across the USA, we can find no meaningful signal of temperature on transmission. This provides two important, and timely, insights for policy-makers: summer weather is no substitute for mitigation, and policy can prevent transmission in the winter. At the coarse scale of USA states, population density is a greater driver of transmission intensity than temperature in our epidemiological modelling (log_10_(population density) ≈ 1.4× larger scaled coefficient than temperature). It should be considered whether thresholds for adaptive and/or intermittent lockdown might be more precautionary (*i.e*., lower) in colder, more densely-populated regions. However, we strongly suggest that this should neither be in order to allow other regions to actively relax restrictions, nor conducted without further examination of finer-scale disease dynamics. When making decisions about the relative importance of climate and population density, it is important to account for the magnitude of variation in the two variables. Temperature varies widely across the US, and that differences in transmission rates between states (contrast, for example, Minnesota and Florida in figure 2) may vary due to climate does not imply that more modest climate differences within a state drive differences through time or across space. Regardless, our analysis is too spatially coarse to address such variation. Even quite large variations in climate are more straightforward to mitigate than population density differences (figure 3), and so we suggest that regions with higher population density should continue to be monitored carefully. Finally, we emphasise that population density and temperature are well-known to be strongly correlated across USA states (see also figure 2); this does not affect our model fitting of coefficient estimates, but it does affect their interpretation. A more densely-populated state is also likely to be warmer, and so we suggest that both factors (and others, such as mobility) should be taken into account when trying to a prion estimate a region’s transmission rate.

**Figure 3:**
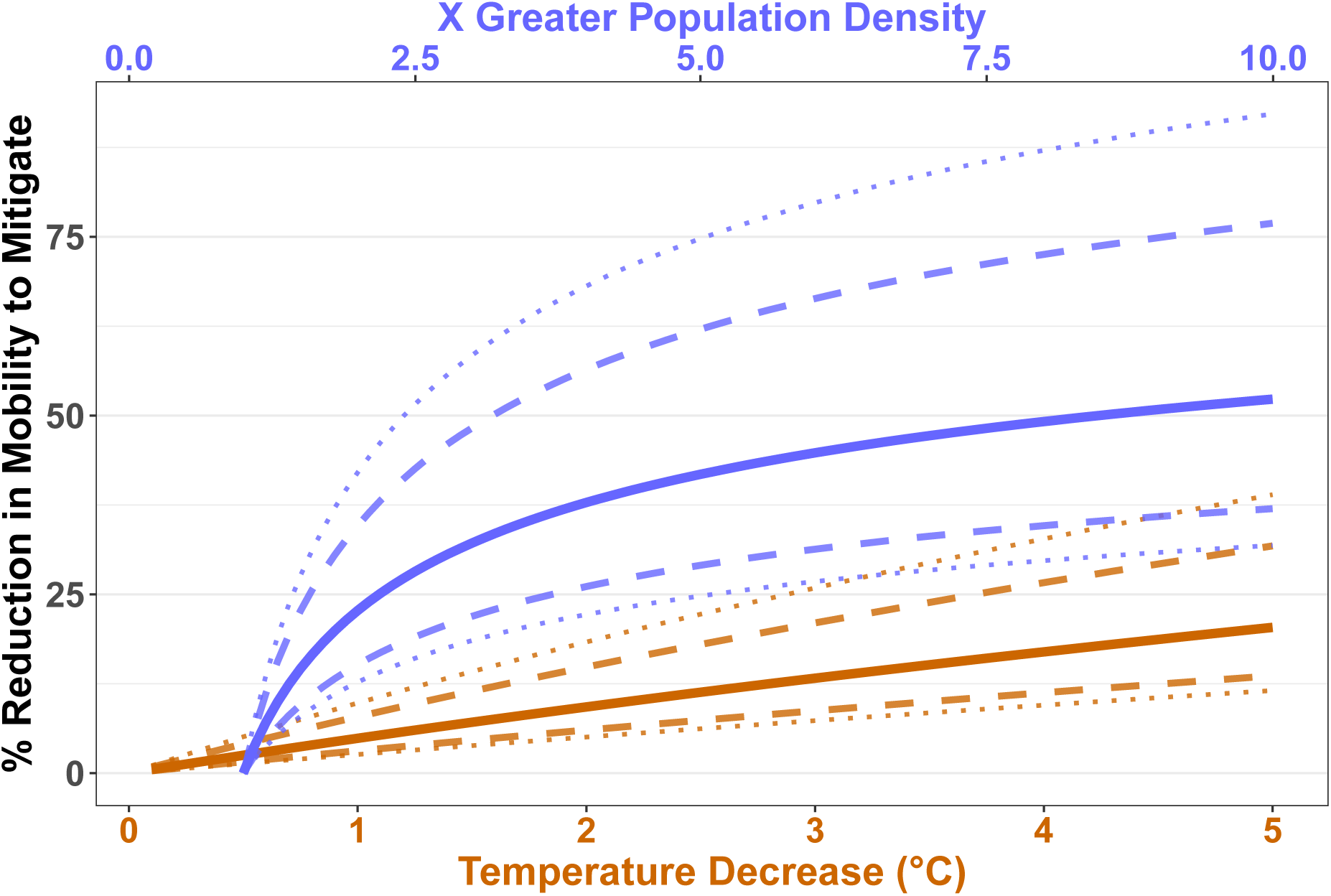
Average mobility reductions required to mitigate differences in population density and temperature. This figure shows the percent reduction in average mobility (measuring retail, recreation, grocery, pharmacy, and workplace trips) needed to compensate for a given temperature (brown) or population density (blue) driven increase in *R_t_*. These calculations assume a ‘background’ *R*_0_ of 1 and a baseline ‘background’ mobility (defined as ‘0’ by Google^36^). Solid lines represent the median mobility reduction required, dashed and dotted lines the 75% and 90% posterior credibility intervals respectively.

These results have strong implications for modellers considering the potential impacts of seasonality on the virus. Such work has already considered the role that seasonality might play by assuming responses of structurally similar and/or related diseases are adequate proxies for SARS-CoV-2^18^. These assumptions are broadly correct, but here we parameterise and quantify the magnitude of this effect for SARS-CoV-2. Our findings suggest that previously unexplained variation among regions’ transmission, such as in our independently-estimated *R*_0_ data, can be accounted for by environmental factors. Further, our results support a role for daily temperature changes in transmission, but, we emphasise, do not conflict with other studies suggesting that seasonal forecasting plays a secondary role to mitigation and/or number of susceptible individuals. Such studies^18^ assumed SARS-CoV-2 responds to climate to broadly similar extents that we find here. What our results do suggest, however, is that future forecasting work should consider the use of the environment to enhance predictions of disease spread. In countries such as the USA with continental climates that swing between extremes of heat and cold, we suggest policy-makers should assume that transmission will increase in winter (and potentially autumn/fall). The timing of the seasons are broadly predictable, and so this is an area in which policy could be proactive, not reactive.

## Conclusion

There is no single cause of, or solution to, the current COVID-19 pandemic, and all drivers must be placed in perspective. Here we suggest that both environment, and daily weather, may play a role in the transmission of SARS-CoV-2. The major driver of transmission, and our best method of controlling it, is public policy, as this and many other studies have shown^40,41^. The role of environment in transmission has become controversial, in part because of the application of models to case prevalence, rather than fundamental epidemiological parameters such as *R* that we model here. We call for more researchers to work directly with epidemiologists to identify those components of epidemiological modelling that can be informed by outside specialists’ areas of expertise. By working together, we can efficiently increase epidemiological research capacity to better combat and control this pandemic.

## Data Availability

o new data are released as part of this project. Code to reproduce the full analysis pipeline is available from our GitHub repository (https://www.github.com/pearselab/tyrell).

## Methods

We explored the association between environmental covariates and SARS-CoV-2 transmission intensity using two approaches. First, we took existing state-level estimates of *R*_0_ and during-lockdown *R_t_* for the USA^35^, and regressed them against environmental data in order to test for potential pre- and during-lockdown patterns. In the second approach, we modified and fitted the existing semi-mechanistic epidemiological model used to generate the *R*_0_ and *R_t_* estimates above, and fitted it to the observed death time series whilst explicitly incorporating the effects of the most important aspects of environment (temperature and population density) on the virus. This second model makes use of daily weather observations and provides a rigorous framework to quantify the drivers of SARS-CoV-2 transmission across the USA. The first approach mitigates potential biases arising from the autocorrelation of the initiation of lockdown and the cessation of winter in the USA in the second approach, since our independent regression focuses on initial transmission (*i.e*., *R*_0_). Below, all software packages given in *italics* are *R* packages (version 3.6.3)^42^ unless otherwise specified. Code to reproduce our analyses, download source data, and update models with new data as it becomes available, are given in both the supplementary materials and at our team’s *GitHub* repository (https://www.github.com/pearselab/tyrell).

### Data collection

We collated global population density data from the Center for International Earth Science Information Network (CIESIN)^43^, and hourly temperature (*T*), relative humidity (*RH*) and surface UV radiation (in J m^-2^) estimates for 2020 from the Copernicus Climate Change Service^44^. All the above data were at the same spatial resolution of 0.25×0.25°. The amount of water vapour air can hold increases with temperature and, since in other viruses the absolute humidity (*AH*) of air can drive transmission more than relative saturation^37,45^, we calculated absolute humidity from our data using the the Clausius-Clapeyron relation and the ideal gas law^22,45^:

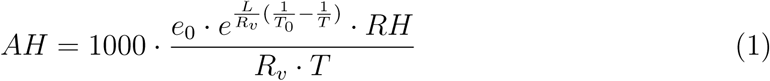

Where *AH* (g m^-3^) is the absolute humidity, *T* (K) the temperature in a given cell, *RH* the relative humidity in a given cell (expressed as a percentage), e_0_ the saturation vapor pressure (6.11mb) at reference temperature *T*_0_ (which we set as 273.15K), L the latent heat of evaporation for water (2257 kJ kg^-1^), and *R_v_* the gas constant for water vapour (461.53 J kg^-1^K^-1^).

We used the Climate Data Operators program^46^ to compute daily means for each of our climate variables. Finally, we averaged the value of each covariate (median) across the state-level administrative units given by GADM shapefiles^47^ (the 50 USA states, plus Washington DC).

### Independent validation of the impact of environment on *R*_0_

To validate the impact of the environment on *R*_0_ we used an existing dataset of SARS-CoV-2 transmission rate estimates for each of the states of the USA^35^. We used the basic reproduction estimates (*R*_0_, before the implementation of any non-pharmaceutical interventions) as a fundamental measure of virus transmissibility in each state.

In these data, *R*_0_ is estimated as *R_t_*_=0_ where *t* = 0 occurs 30 days prior to the first 10 cumulative deaths recorded for each state^32,35^. The date upon which *R*_0_ is estimated therefore differs between states. To account for these temporal differences, we took the means of our daily climate variables across the 14 days prior to *t* = 0 for each state as an approximation for the conditions under which each population first experienced COVID-19. To test the impact of the environment on *R*_0_, we performed multiple linear regression on *R*_0_ with temperature, absolute humidity, UV radiation, and population density as predictors. To compare environmental effects on the reproduction number under mobility restriction measures *(i.e*., lockdown), we took the average (mean) *R_t_* across the 14 days following a state-wide stay-at-home mandate and regressed these against the environmental predictors averaged across the same time period. We used 14 days again here for consistency with our environmental comparison to *R*_0_. Although mobility restrictions may differ in magnitude between states, these effects are incorporated into the estimates for the *R_t_* parameter. In 7 states (Arkansas, Iowa, North Dakota, Nebraska, Oklahoma, South Dakota, and Wyoming) no state-wide stay-at-home mandate was declared. In a further 4 states (Alaska, Hawaii, Montana, and Utah), *t* = 0 occurred after non-pharmaceutical interventions had already been instated. These 11 states were therefore excluded from the independent validation analyses.

### Integrative modelling of the impact of environment on SARS-CoV-2 transmission

To further assess the potential impact of environment on SARS-CoV-2 transmission, we modified the semi-mechanistic Bayesian model^35^ that generated the *R*_0_ and *R_t_* estimates used above to incorporate both population density and daily temperature (the best fitting climate variable; see results), as follows:

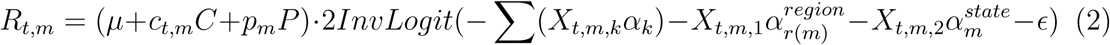

Where *μ* captures overall transmission common to all states, *c* the coefficient for temperature *(C_t,m_;* in degrees C) at time (*t*) in state *m, p* the coefficient for population density *(P_m_)* of state *m* (log_10_-transformed people per km^2^). We standardised *c_t,m_* and *p_m_* to have a mean of 0 and standard deviation of 1, in order to make their absolute magnitudes measures of the relative importance of each term and thus facilitate their comparison. We placed strong, conservative priors on these new model terms, specifically:

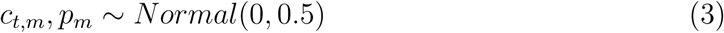

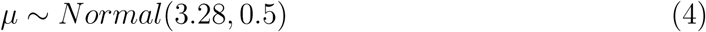

For *μ*, this is the same as the prior used in the original (non-climate) model^35^ (but see our caveat below about this term). The other terms are unchanged from their original definitions given in Unwin et al.^35^, and we briefly describe them below. *InvLogit* is the inverse logit transformation applied to a series of hierarchically-nested terms *(α_k_*, 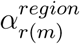, and 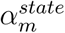) multiplied by Google mobility data^36^ *(X_t,m,k_*) with a weekly *AR(2)* autocorrelated error term for each state (e; see Unwin et al.^35^ for more details). *X_t,m,k_* are three US-wide measures of the impact of changing mobility across states (a daily proxy for lockdown intensity) on ‘average’ across retail and recreation, grocery and pharmacy, and workplace trips (*X_t,m,_*_1_), in ‘residential’ areas (*X_t,m,_*_2_), and using public ‘transit’ *(X_t,m,_*_3_). We focus on the vector *α_k_*, whose three entries assess the impact of mobility comparably across the country (and thus are each analogous to *c* and *p*). The terms 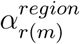 and 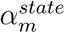 address differences in average mobility across eight broad geographic regions [the Great Lakes, Great Plains, Rocky Mountains, Northeast Corridor, Pacific North-West, South Atlantic, Southern Appalachia, and the South (‘TOLA’); indexed by *r* (*m*)] and for transit across individual states (*m*), respectively. While we attempted to address comparable hierarchically-nested temperature responses in this model, we felt the correlation between changes in *X_t m k_* and *c_t,m_* were inducing fitting problems and so opted for a model simpler (and so more conservative) in its novel components. In this model formulation, temperature and population density essentially contribute to a latent transmission rate, which is then mediated by the mobility terms to produce the realised *R_t_*. Although an interaction between mobility and environment (as found in our regression modelling, see Results) is not explicitly modelled, this formulation produces results analogous that finding, *i.e*. when mobility reductions are high (“lockdown”), environment has little effect on the realised *R_t_* (see supplementary figure S2).

We emphasise that the model presented here differs from the original model by fitting a common *μ* across all states, instead of allowing each state to have a different baseline *μ* that was hierarchically drawn from a common parameter (itself termed *μ* in Unwin et al.^35^). This difference ensures identifiability of our model parameters, since the (latent, and hierarchically pooled) state-wise means are strongly driven by both population density and environment that are now included in the model (see results). Our model, which was directly adapted from the code in Unwin et al.^35^, was fit using *rstan*^48^ with 5 independent chains (each with 3,000 total iterations and a warm-up of 1,000). Full model coefficients and outputs are given in the supplementary materials (supplementary table S6); posterior predictive checks were performed to ensure that the predicted *R_t_* values for each state through time were realistic and sensible and all chains had mixed and converged.

## Data and Code Availability Statement

No new data are released as part of this project. Code to reproduce the full analysis pipeline is available from our *GitHub* repository (https://www.github.com/pearselab/tyrell).

## Acknowledgements

This work was funded by a Natural Environment Research Council grant NE/V009710/1. WDP and the Pearse Lab are also funded by National Science Foundation grants ABI-1759965 and EF-1802605.

